# Proteomics Uncovers Immunosuppression in COVID-19 Patients with Long Disease Course

**DOI:** 10.1101/2020.06.14.20131078

**Authors:** Shaohua Tang, Rui Sun, Qi Xiao, Tingting Mao, Weigang Ge, Chongquan Huang, Meng Luo, Liujia Qian, Hao Chen, Qiushi Zhang, Sainan Li, Wei Liu, Shufei Li, Xueqin Xu, Huanzheng Li, Lianpeng Wu, Jianyi Dai, Huanhuan Gao, Lu Li, Tian Lu, Xiao Liang, Xue Cai, Guan Ruan, Kexin Liu, Fei Xu, Yan Li, Yi Zhu, Jianping Huang, Tiannan Guo

## Abstract

Little is known regarding why a subset of COVID-19 patients exhibited prolonged positivity of SARS-CoV-2 infection. Here, we studied the sera proteomic dynamics in 37 COVID-19 patients over nine weeks, quantifying 2700 proteins with high quality. Remarkably, we found that during the first three weeks since disease onset, while clinical symptoms and outcome were indistinguishable, patients with prolonged disease course displayed characteristic immunological responses including enhanced Natural Killer cell-mediated innate immunity and regulatory T cell-mediated immunosuppression. We further showed that it is possible to predict the length of disease course using machine learning based on blood protein levels during the first three weeks. Validation in an independent cohort achieved an accuracy of 82%. In summary, this study presents a rich serum proteomic resource to understand host responses in COVID-19 patients and identifies characteristic Treg-mediated immunosuppression in patients with prolonged disease course, nominating new therapeutic target and diagnosis strategy.

## INTRODUCTION

COVID-19 caused by the SARS-CoV-2 virus is an ongoing pandemic spreading all over the world. About 80% of COVID-19 patients exhibit mild clinical symptoms including fever and cough, while ∼20% of patients deteriorate to life-threatening severe or critical conditions (Wu and McGoogan, 2020). Multiple studies have investigated the clinical course and pathogenesis (To et al., 2020; Yang et al., 2020; Zhou et al., 2020) and molecular changes (Messner, 2020; Shen, 2020; Wu et al., 2020) of severe or critical cases.

What remains largely unknown is the substantial diversity of COVID-19 patients in terms of disease course. In a recent report, the median interval of COVID-19 patients from onset to clinical recovery was 20 days, with the longest disease course of 37 days (Zhou et al., 2020). According to our clinical treatment of 37 COVID-19 patients, some patients exhibited surprisingly prolonged virus infection. The longest disease course we have observed thus far is 107 days (P33, a 55-year-old male) as of 17 May 2020. Little is known about the causes of persistent virus infection and how he could be effectively treated.

Here we report a longitudinal and in-depth proteomic profiling of 37 COVID-19 patients with short and long disease courses. We analyzed the sera proteomes collected over nine weeks for both patients with short disease course (SC) who turned negative for the virus infection in less than 22 days, and patients with long disease course (LC) who remained positive for 22 days or longer. We have also analyzed the sera proteomes of 35 non-COVID-19 patients with flu-like symptoms as controls. 2700 proteins were quantified with high quality, offering a rich resource to understand the dynamic disease course of COVID-19 patients in response to virus infection. Our data showed characteristic immune responses between the SC and LC patient groups. Remarkably, the data suggest regulatory T cell (Treg)-mediated immunosuppression was specifically activated in LC patients. We also showed that it is possible to predict the disease course of COVID-19 patients by measuring the characteristic protein levels from blood samples collected in the first three weeks since disease onset using a machine learning model. This study not only presents a rich resource to study the COVID-19 host responses, but also nominates novel therapeutic and diagnostic strategies for COVID-19 patients with prolonged infection positivity.

## RESULTS AND DISCUSSION

### Patients, samples and proteomics

We procured 37 COVID-19 patients in this study of which 35 were typical and two were severe cases (Table 1, Table S1), including 19 males and 18 females, with a median age of 48 years. The median duration of disease in this cohort, as defined in Methods, was 22 days. These patients were divided into two groups based on the length of their disease course (Figure 1). The SC group comprised 17 patients with a disease course shorter than 22 days. The other 20 patients were defined as LC group. Interestingly, one of the severe cases was classified as SC, while the other severe case was in the LC group (Figure 1).

**Table 1.** Baseline and progression of CVDTSA cohort.

| Baseline Characteristic | non-COVID-19<br>(N=35) | COVID-19 SC<br>(N=17) | COVID-19 LC<br>(N=20) |
| --- | --- | --- | --- |
| **Gender — no. <sup>a</sup>(%)** |  |  |  |
| male | 17 (53.1) | 8 (47.1) | 11 (55.0) |
| female | 15 (47.9) | 9 (52.9) | 9 (45.0) |
| **Age — yr <sup>b</sup>** |  |  |  |
| mean ± SD | 43.0 ± 18.0 | 43.2 ± 10.5 | 49.0 ± 15.4 |
| median (IQR) | 37.5<br>(28.8-56.0) | 44<br>(35.0-50.0) | 50<br>(42.8-55.5) |
| range | 18-80 | 27-67 | 2-84 |
| **Symptoms —no. (%)** |  |  |  |
| fever | 28 (87.6) | 10 (58.8) | 14 (70.0) |
| cough | 2 (6.3) | 11 (64.7) | 15 (75.0) |
| diarrhea | 1 (3.1) | 10 (58.8) | 8 (40.0) |
| fatigue | 1 (3.1) | 8 (47.1) | 7 (35.0) |
| **Comorbidities — no. (%)** |  |  |  |
| hypertension |  | 2 (11.8) | 6 (30.0) |
| diabetes |  | 0 (0.0) | 3 (15.0) |
| hepatitis B |  | 2 (11.8) | 0 (0.0) |
| coronary sclerosis |  | 0 (0.0) | 3 (15.0) |
| gastrohelcosis |  | 1 (5.9) | 0 (0.0) |
| psoatic strain |  | 0 (0.0) | 1 (5.0) |
| chronic gynecologic inflammation |  | 1 (5.9) | 0 (0.0) |
| gout |  | 0 (0.0) | 1 (5.0) |
| asthma |  | 0 (0.0) | 1 (5.0) |
| **Treatment— no. (%)** |  |  |  |
| lopinavir and ritonavir |  | 17 (100.0) | 19 (95.0) |
| Atomized interferon |  | 17 (100.0) | 20 (100.0) |
| Arbidol |  | 7 (41.2) | 12 (60.0) |
| Lianhuaqingwen (Chinese traditional medicine) |  | 15 (88.2) | 16 (80.0) |
| Ribavirin |  | 0 (0.0) | 3 (15.0) |
| Hydroxychloroquine |  | 1 (5.9) | 3 (15.0) |

<sup>a</sup> no.: number.<sup>b</sup> yr.: year

**Figure 1.**
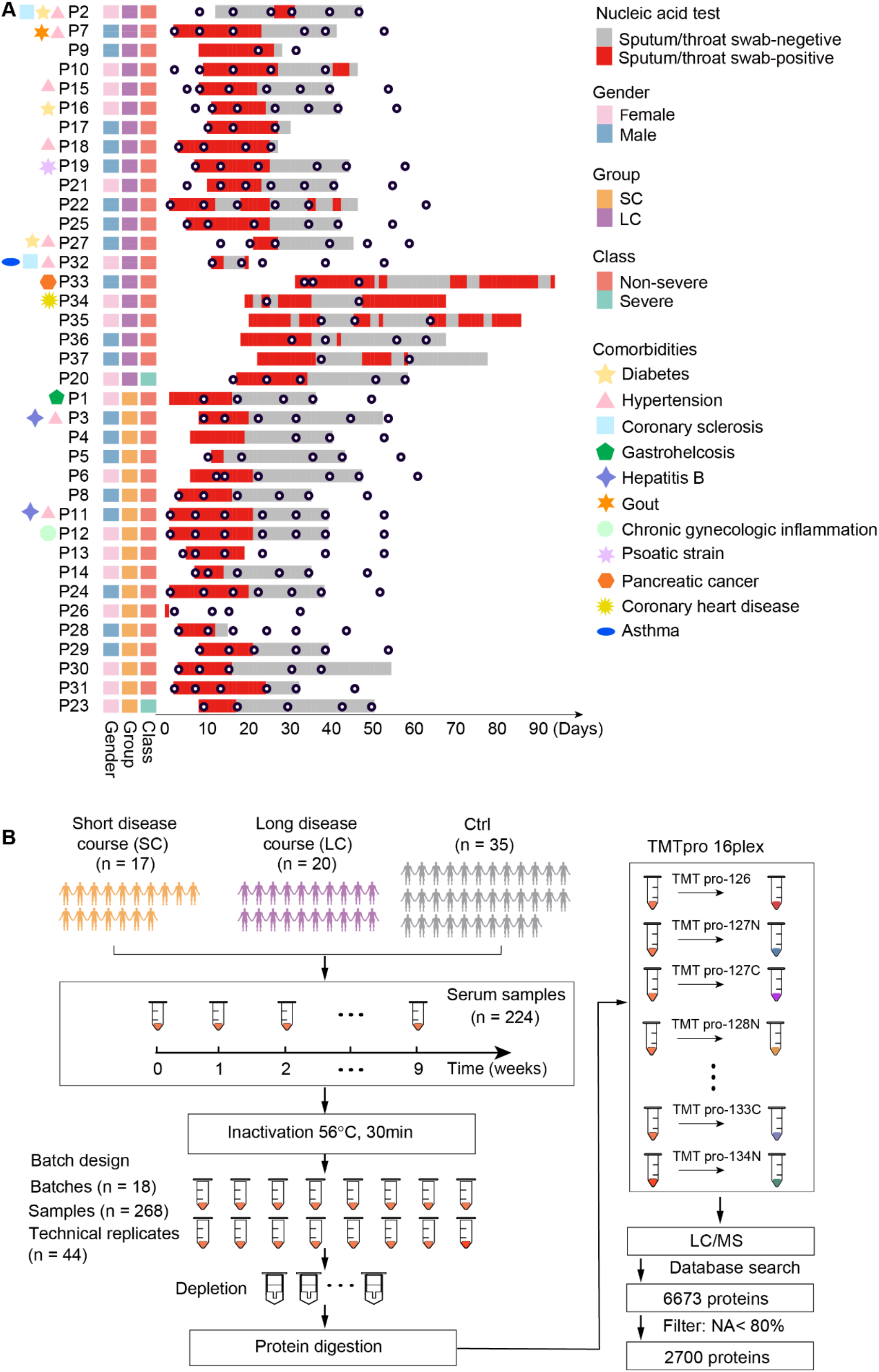
Patients, samples and study workflow. **(A)** Baseline characteristics of the study cohort. The y-axis shows the patient ID, and the x-axis displays the length of disease course from onset. The 37 patients are separated into SC (17 patients) and LC (20 patients) groups. Other important information including the virus nucleic acid test results (sputum/throat swab-positive/-negative), gender, severity, comorbidity baseline, etc., are shown in the right panel of the figure. The black dots indicate sampling time. More details are provided in Table S1. **(B)** Workflow for TMT based proteomic analysis in this study. 37 COVID-19 patients and 32 control (Ctrl) patients were included. In total, 224 sera samples were collected and 268 peptide samples including 44 technical replicates were analyzed by TMT 16plex-based quantitative proteomics.

These two groups (SC and LC) were indistinguishable based on clinical symptoms. The median age of SC patients was 43.2 years (range, 27-67 years) for SC patients, and 49.0 years (range, 2-84 years) for LC patients. The Wilcox t-test showed no significant age difference between the SC and LC patient groups (Figure S1A). Clinical treatment of both groups of patients was not significantly different by Fisher’s exact test (Figure S1B, Table 1), indicating that the different lengths of disease course were unlikely due to existing therapeutics. Routine blood test data for the two groups showed no significant difference either (Figure S1C). We analyzed an additional 35 non-COVID-19 patients with flu-like symptoms as controls (Table 1, Table S1, and Figure 1B).

We performed in-depth profiling for sera proteome collected from each patient over nine weeks using TMTpro 16plex-based data-dependent acquisition mass spectrometry (Thompson et al., 2019). 14 high-abundance blood proteins were largely depleted to enhance the ability to measure low-abundance proteins. Each TMT-labeled peptide mixture was fractionated into 26-30 fractions for comprehensive discovery proteomic analysis (Figure 1A). A total of 224 depleted serum samples were analyzed using TMT-based proteomics (Figure 1). Samples were randomly distributed into 18 batches. We also analyzed 44 technical replicates for randomly selected samples in each batch (Figure S2A). Each pool of TMT-labeled peptides was further fractionated into 26-30 fractions using high-pH reverse phase liquid chromatography (RPLC) and analyzed by LC-MS/MS using a 60-min RPLC gradient. The median coefficient of variance (CV) among MS/MS technical replicates was 0.19 (Figure S2B). After excluding proteins with missing values in over 80% of samples, we presented quantitative levels of 2700 proteins across 268 sera samples with relatively high degrees of quantitative accuracy (Table S2A).

### Complex immunity in the LC patients

To investigate host response differences in the serum proteome data set from the SC and LC patients, we performed a pairwise comparison of the proteins quantified in these two groups at each week over nine weeks. We then combined the regulated proteins at each week, leading to the identification of 921 dysregulated proteins (Table S2B), of which 405 proteins were functionally annotated in the UniProt database. We firstly used only GO biological process, KEGG and Reactome pathway enrichment using 405 proteins, and found that most pathways were related to immunity and metabolism (Figure S3).

Generally, our data showed that the first week is the inflammatory response period, featured by activation of redox and amino acid metabolic processes. In the following three weeks, more pathways involved in adaptive immunity and glycometabolism were activated. Interestingly, some sex hormone associated pathways were also enriched, such as fertilization and response to estradiol. From fifth to ninth week, the two patient groups displayed difference in pathways involved in tissue remodelling and repair, apoptosis and sulfur compound catabolic process.

We then focused on the immunity-related pathways at each time point as shown in Figure 2A. For SC patients, they showed upregulated platelet degranulation in the first week, followed by activation of antigen presentation in the following two weeks. Leukocyte degranulation was activated in the third week. In the sixth week, SC patients showed upregulation of serum proteins involved in tissue remodeling (Figure 2A) as signs of recovery. LC patients displayed much more complex serum proteome dynamics. During the first week, both platelet degranulation and upregulation of Treg vs Teff were activated. Activation of antigen presentation occurred in the second week. The complement system was also activated from the second week.

**Figure 2.**
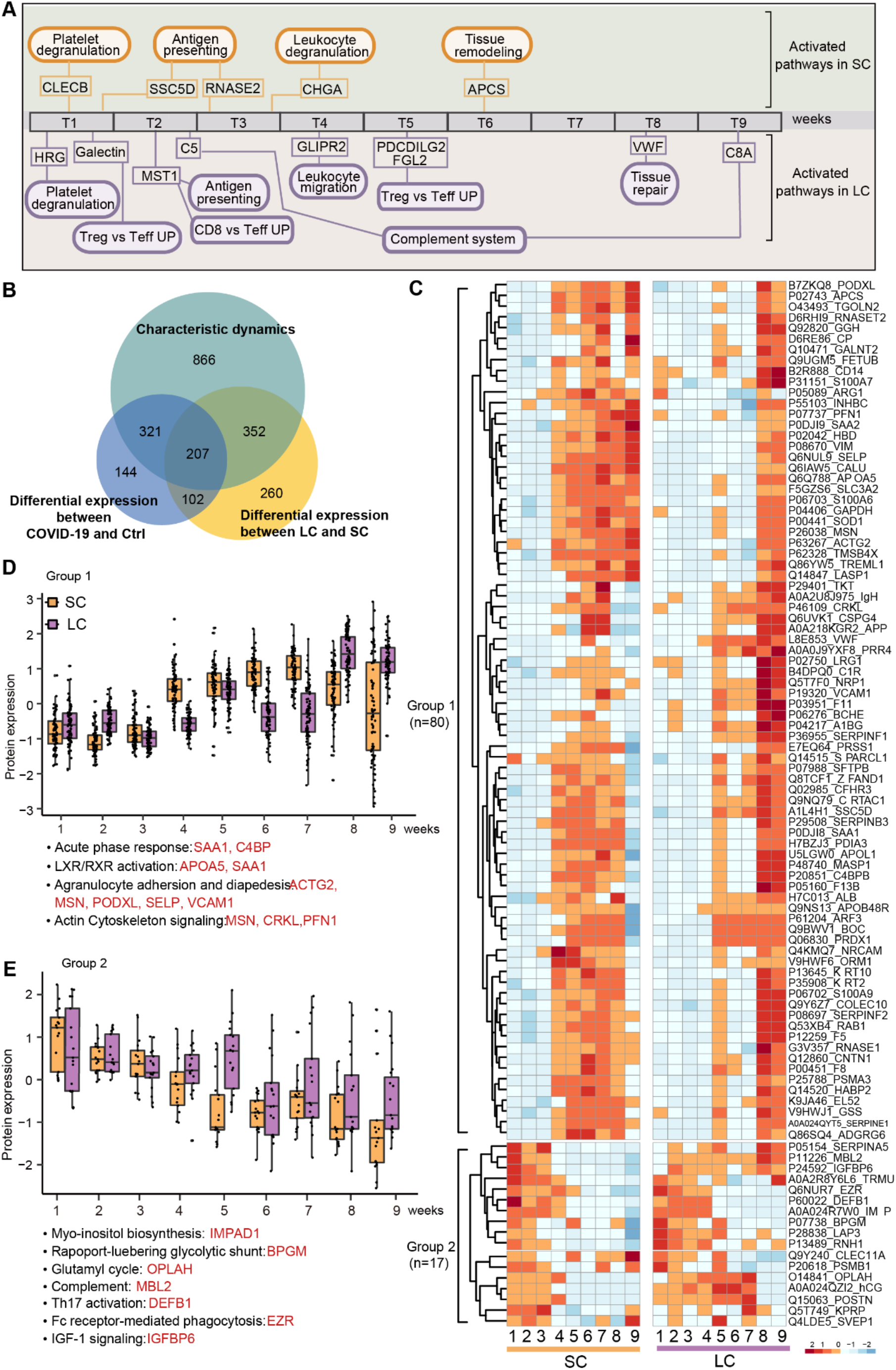
Complex immunity in the LC patients. **(A)** Summary of enriched immune-related pathways in LC and SC patients across the disease course. Representative proteins involved in these pathways are also shown. **(B)** Venn diagram of differentially expressed proteins between the SC and LC groups over nine time points and specifically regulated in COVID-19 sera. **(C)** Heatmap of 97 dysregulated proteins in the SC and LC groups over nine time points. **(D-E)** Expression of protein group 1 and 2 across nine time points. The y-axis stands for the relative protein expression normalized by z-score. The pathways and representative participating proteins are shown below the boxplots.

Interestingly, the regulatory T cell (Treg) activation pathway was enriched in the first two weeks and the fifth week, suggesting immunosuppression occurred early in these patients. From the fourth week, leukocyte migration was enriched. Tissue repair was enriched only in the eighth week, followed by the complement system in the ninth week (Figure 2A). These data suggest that immunological responses during the first three weeks may be critical in determining the length of the disease course. During this period, both innate and adaptive immunity were activated in SC patients, while in contrast, LC patients showed signs of Treg-mediated immunosuppression in addition to innate and adaptive immunity.

Our data also underscored the proteins involved in the above-mentioned pathways as shown in Figure S3. In SC patients, a secreted protein, the soluble scavenger receptor cysteine-rich domain-containing protein (SSC5D) was highly expressed in the second week (Figure S4). SSC5D is highly expressed on monocytes and activated lymphocytes rather than naïve immune cells, and mediates both innate and adaptive immunity (Goncalves et al., 2009), suggesting activated immune responses to virus infection. In the third week, another upregulated protein in SC patient sera was non-secretory ribonuclease (RNASE2), a chemoattractant for dendritic cells. RNASE2 is reported to participate in antigen presentation (Yang et al., 2003), and may play a role in the adaptive immunity against SARS-CoV-2 (Figure S4). In the fourth week, a pro-inflammatory factor, chromogranin-A (CHGA), was upregulated in the sera in SC patients. CHGA highly expressed on the secretory cells to regulate catecholamine secretion which is associated with serum CHGA (di Comite et al., 2006) and is reported to activate pro-inflammatory M1 macrophages and inhibit anti-inflammatory M2 macrophages (Eissa et al., 2018). In the sixth week, serum amyloid P-component (APCS) was upregulated in SC patient sera (Figure S4), suggesting clearance of damaged circulating cells and enhanced angiogenesis.

In LC patients, positive regulators for immune response were found to be activated. Hepatocyte growth factor-like protein (MST1) was upregulated in the second week (Figure 2A). MST1 is required for thymocyte migration and antigen recognition (Ueda et al., 2012). Besides, Golgi-associated plant pathogenesis related protein 2 (GLIPR2), associated with the migration of T and B lymphocytes, was up-regulated in the fourth week (Figure S4). In the following week, we observed upregulation of two immunosuppressive factors, namely fibroleukin (FGL2) and programmed cell death 1 ligand 2 (PDCD1LG2). FGL2 negatively regulates dendritic cell antigen presentation and T cell differentiation, especially the secreted FGL2 (Hu et al., 2016). It was upregulated in LC patients and further increased during recovery from COVID-19 (Figure S4). PDCD1LG2 negatively regulates T cell activation (Brown et al., 2003), and followed similar dynamics over the disease course to FGL2 (Figure S4). In the eighth week, tissue remodeling associated proteins, such as von Willebrand factor (VWF), were upregulated in the LC group, suggesting recovery from COVID-19. Besides, our data show that the complement system may play important roles in LC patients during the entire disease course. In the second week, the most enriched pathway involving the upregulation of C5 was the complement cascade (Figure 2A). In the last week, C8A and other complement system proteins remained upregulated (Figure S4). Complement activation was reported in our previous study of COVID-19 sera (Shen, 2020). The complement cascade could also contribute to immunosuppression (Kemper et al., 2003) and might be a potential therapeutic target for COVID-19 (Cao, 2020).

Altogether, negative regulator factors and innate immune performed more active might result in longer disease course.

### Characteristic protein dynamics in the LC patients

Next, we investigated whether the differentially expressed proteins between SC and LC patients were unique to COVID-19 disease by making comparison between COVID-19 and non-COVID-19 patients with flu-like symptoms (Ctrl, Figure 1B). We first analyzed the temporal dynamics of the 2700 proteins using Mfuzz (Kumar and Mattias, 2007) for SC and LC patients. 1746 proteins, grouped in eight clusters in the SC and LC patient groups (Table S2C), were consistently dysregulated over time compared to the control group (Figure S5A-B). Four characteristic protein clusters were enriched in the two patient groups (Figure S5C). In the meantime, 774 proteins were identified to be differentially expressed in COVID-19 patients compared to the Ctrl group (Table S2D). By overlapping the two above-mentioned protein shortlists with the 921 dysregulated proteins between SC and LC patients, our data were narrowed down to 207 proteins which fulfill the following three criteria: a) differentially expressed between SC and LC; b) consistently up- or down-regulated over time; c) COVID-19 disease specific (Figure 2B). Out of 207 proteins, 97 proteins were functionally annotated by the Ingenuity Pathway Analysis (IPA) software tool (Table S2E).

These proteins were segregated into two major groups based on their expression across all nine time points during the disease course (Figure 2C). The first group of 80 proteins showed increased expression over time. SC patients exhibited earlier elevation of these proteins, while LC patients exhibited delayed increase (Figure 2C). Four major pathways were enriched in this protein group, namely acute phase response, LXR/RXR activation, agranulocyte adhesion and diapedesis, and actin cytoskeleton signaling (Figure 2D). Also, the coagulation and complement systems were also enriched in this protein group. Acute phase response signaling, coagulation and complement systems are associated with innate immunity, while actin cytoskeleton signaling is associated with leukocyte migration in adaptive immunity.

The second group of 17 proteins showed decreasing expression over time. SC patients showed an earlier decrease than LC patients (Figure 2E). These proteins were mapped to enriched pathways including metabolic processes such as myo-inositol biosynthesis, glycolytic shunt and glutamyl cycle (Figure 2E).

Next, we calculated the median expression level of the two groups of proteins and present them in boxplots (Figure 2D, 2E). For the first group of proteins, generally SC patients showed lower protein expression than LC patients in the first three weeks, but the expression was dramatically elevated from the fourth week and maintained thereafter at a high level for several weeks (Figure 3D). This agrees with clinical data that all these SC patients tested negative for SARS-CoV-2 nucleic acid after the third week. In contrast, LC patients showed a higher degree of longitudinal diversity in the trends of expression of these proteins, probably partly due to their variable clinical recovery trajectories. Expression of the first group of proteins in the first four weeks was relatively stable but dramatically increased in the fifth week, followed by a decrease from the sixth to the seventh week, and a second rise in the eighth week (Figure 2D).

**Figure 3.**
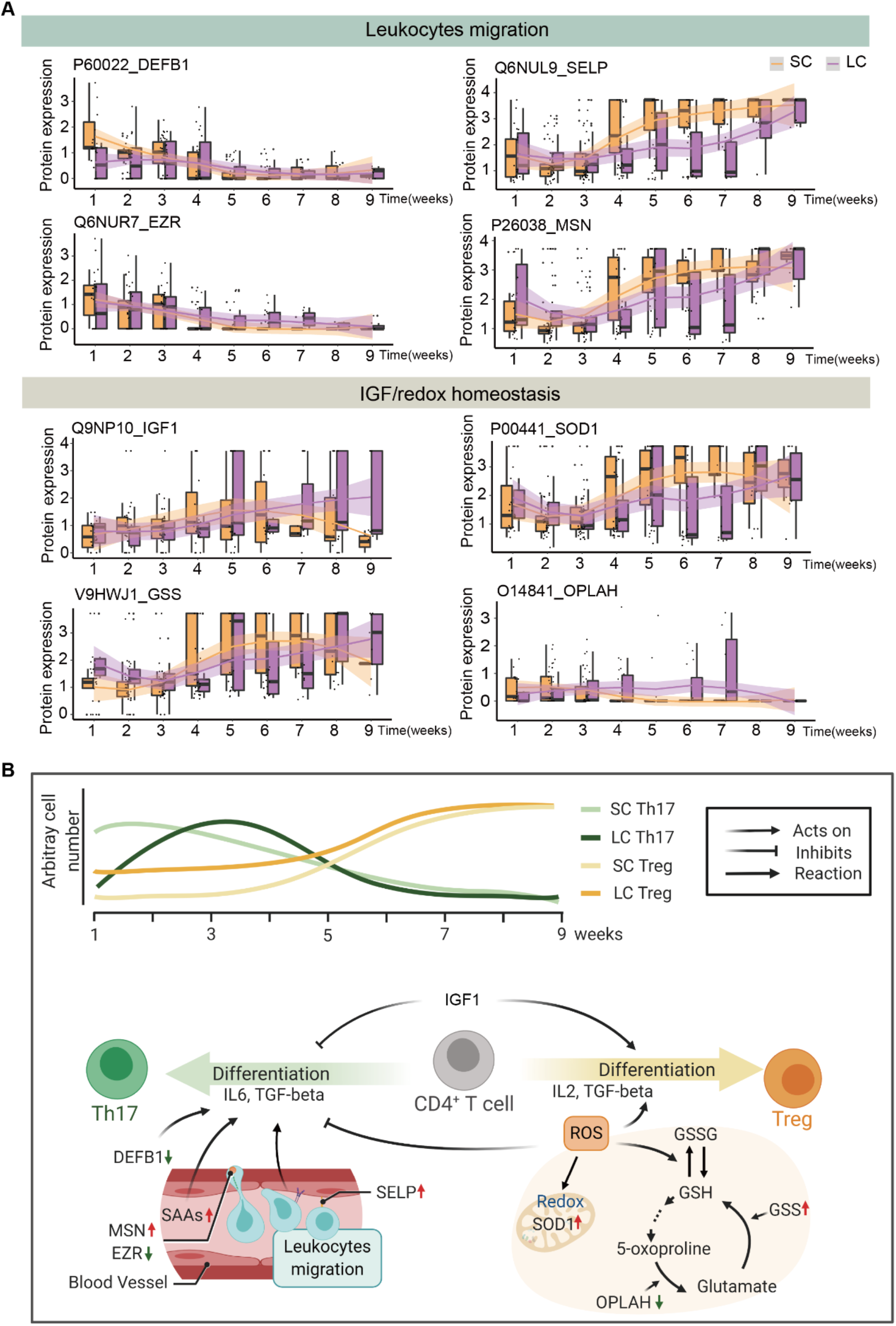
Characteristic proteins and pathways in LC patients. **(A)** Representative proteins involved in leukocyte migration and IGF/redox homeostasis are plotted over time. **(B)** Diagram depicting that major regulated pathways in the LC patients including leukocytes migration and IGF/redox homeostasis jointly modulate Treg-mediated immunosuppression.

Interestingly, the expression of the second group of proteins displayed an opposite trend compared to the first group (Figure 2E). Protein expression of the SC patients started to decrease during the 4^th^ week and continued to decrease until the last time point; while the LC patients showed high protein expression level till the fifth week, with a shallow decrease beginning in the sixth week (Figure 2E).

### Dysregulated leukocyte migration and IFG/redox homeostasis

Pathway analysis of the two protein groups mentioned above highlighted leukocyte migration and IGF/redox homeostasis (Figure 3).

Representative proteins related to leukocyte migration included beta-defensin 1 (DEFB1), selectin P (SELP), ezrin (EZR) and moesin (MSN) (Figure 3A). DEFB1 is a ligand for the chemokine receptor CCR6 and a positive upstream regulator of Th17 (Wojtowicz et al., 2015). SC patients exhibited high expression of DEFB1 in the first three weeks, but it decreased from the fourth week when patients became negative in the nucleic acid test (Figure 3A). In contrast, LC patients exhibited a delayed increase of this protein until the third week (Figure 3A), suggesting delayed activation of Th17 in LC patients. Leukocyte extravasation occurs in two sequential steps, among which SELP participates in the first process of leukocyte adhesion to the endothelial surface (Vicente-Manzanares and Sanchez-Madrid, 2004). In SC patients, expression of SELP was relatively low in the first three weeks, and then increased significantly in the fourth week and maintained at a high level until the ninth week (Figure 3A). SELP increase in LC patients, however, was much shallower, and peaked as late as the eighth week, in line with their prolonged disease course (Figure 3A). The complex of ezrin-radixin-moesin (ERM) links cytoskeleton with membrane adhesion molecules; however, moesin (MSN) and ezrin exert opposite functions (Ivetic et al., 2002). Our data showed that EZR was maintained at a relatively high level during the first three weeks in SC patients and then decreased thereafter (Figure 3A). Interestingly, in these patients, MSN showed an opposite trend in which expression increased from the fourth week and thereafter was maintained at a relatively high level. In LC patients, the change of ERM components showed a similar pattern but with a delayed transition point (Figure 3A).

In addition to the proteins directly involved in immunity, our data also highlighted metabolic pathways such as the IGF/redox homeostasis pathway which is critical in maintaining the microenvironment for immune responses (Figure 3A). We narrowed our focus to proteins involved in IGF/redox homeostasis namely insulin-like growth factor 1 (IGF1), superoxide dismutase (SOD1), glutathione synthetase (GSS) and 5-oxoprolinase (OPLAH) (Figure 3A). IGF1 promotes the proliferation of Treg cells (Anguela et al., 2013). In SC patients, expression of IGF1 increased from the second to the sixth week, followed by a dramatic decrease in the seventh week, in line with the recovery of these patients (Figure 3A). However, in LC patients, expression of IGF1 slightly upregulated than in SC patients and increased until the eighth week, followed by a slow decline thereafter. The expression of IGF1 is higher in the LC patients than SC patients in the initial and the end of the disease stage (Figure 3A). Infection-induced ROS was reported to accelerate the accumulation of Treg cells, which led to immunosuppression (Yang et al., 2013). ROS may also induce the expression of SOD1 in the mitochondria to reduce ROS as negative feedback (Ma, 2013). Our data showed that the expression of SOD1 peaked earlier in SC patients (fourth week) than that in the LC patients (fifth week, Figure 4A), indicating that clearance of infection-induced ROS in SC patients occurred earlier than in LC patients. GSS and OPLAH are both involved in the glutamyl cycle that maintains redox homeostasis (Forman et al., 2009). In the glutamyl cycle, GSS catalyzes the generation of reduced glutathione (GSH). OPLAH catalyzes the conversion of 5-oxoproline into glutamate and indirectly enhances the consumption of GSH. Our data showed that GSS was upregulated in the fourth week in SC patients, about one week earlier than its increase in LC patients (Figure 3A). In contrast, the downregulation of OPLAH began in the fourth week in SC patients, while in LC patients it did not substantially decrease until the eighth week (Figure 3A). The timely upregulation of GSS and downregulation of OPLAH might contribute to the elimination of infection-induced ROS in SC patients, while their delayed modulation may have contributed to the prolonged disease course of LC patients.

**Figure 4.**
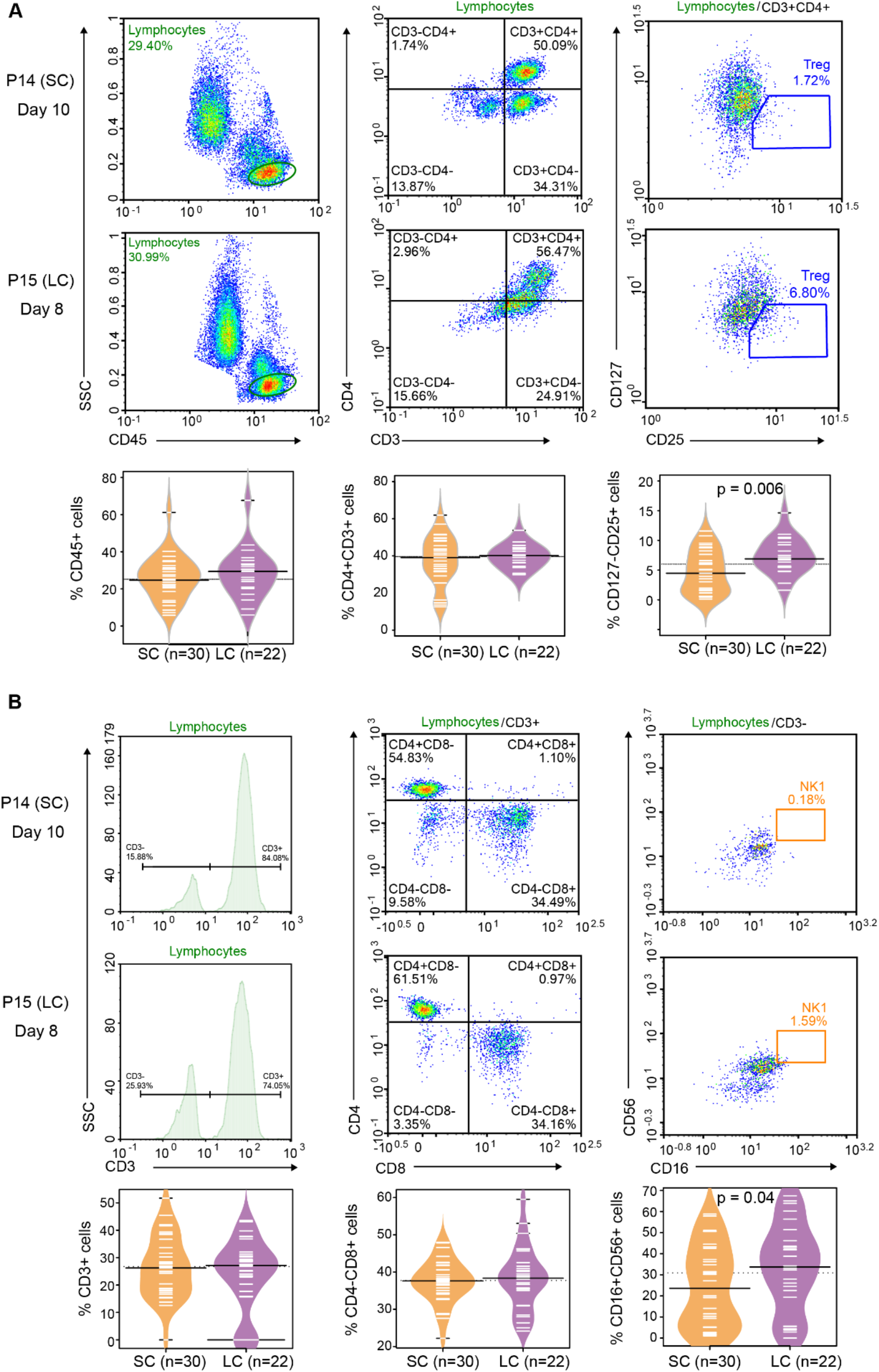
Flow cytometric analysis of immune cells between SC and LC patients. 22 samples from 17 LC patients and 30 samples from 17 SC patients were analyzed. **(A)** Flow cytometric analysis of lymphocytes, CD4+ cells and CD127-CD25+ Treg cells of two representative patients. Bean plots show the comparison between the two groups. **(B)** Flow cytometric analysis of lymphocytes, CD8^+^ cells and CD56+CD16+ NK cells of two representative patients. Bean plots show the comparison between the two groups.

### Treg-mediated immunosuppression and crosstalk with other immune cells in the LC patients

The leukocyte migration and IGF/redox homeostasis pathway may modulate the activities of Th17 and Treg cells (Figure 3B). Th17 cells are a subset of T helper (Th) cells. Th17 cells are differentiated from CD4+ T cells with a pro-inflammatory role upon infection when IL6 is relatively high (Kimura and Kishimoto, 2010). The upregulation of IL-6 has been reported as one of the most critical cytokines in severe COVID-19 patients (Ruan et al., 2020). Our data suggested that activation of Th17 cells may play a role in T cell immunity in COVID-19. Opposite to Th17 cells, Treg cells are induced from CD4+ T cells when IL-6 is relatively low and they suppress immunity (Kimura and Kishimoto, 2010).

Our data showed longitudinal down-regulation of DEFB1, an upstream activator of Th17. LC patients showed a delayed decrease of DEFB1, suggesting delayed activation of Th17. IGF1 promotes Treg cells. A higher level of IGF1 in the LC patients suggested aberrant Treg-mediated immunosuppression in the LC patients (Figure 3A). Besides, infection-induced ROS, regulated by GSS and OPLAH, may also trigger Treg-mediated immunosuppression in LC patients as discussed above.

We next applied flow cytometry to analyze the CD127^-^CD25^+^ Treg cell subpopulation in peripheral blood samples collected from disease onset to the fourth week (SC: 17 patients, 30 samples; LC: 17 patients, 22 samples). The data showed that the number of CD127^-^CD25^+^ Treg cells was significantly higher in LC patients than that in SC patients (p=0.006), while the CD45^+^ lymphocytes (p=0.306) and CD3^+^CD4^+^ T cells (p=0.871) did not show a significant difference (Figure 4A). The data support the hypothesis that CD127^-^CD25^+^ Treg-mediated immunosuppression may be induced in LC patients during the first few weeks. Inhibition of Treg cells could be a potential therapeutic approach for COVID-19 patients with long disease course.

More natural killer (NK) cells, especially the cytotoxic subset (CD3^-^ CD56^+^CD16^+^), were found in LC patients (Figure 4B). NK cells are a typical kind of cells in innate immunity. In the meantime, CD3^-^CD56^-^CD16^-^ cells were found to be decreased in the LC group (Figure S6A-B). These cells are probably B cells, which is further supported by our finding that the antibody level of these patients were also decreased (Huang, 2020). The decrease of B cells may also be supported by the finding that NK cells can inhibit the B cell immunity through reducing neutralizing antibodies (Rydyznski et al., 2015). However, unfortunately we did not have sufficient sample to stain these reduced cells with markers for B cells to confirm their identity.

We then measured the key cytokines and growth factors such as IL-2, IL-4, IL-5, IL-6, IL-10, IL-17A, IFN-γ, and TNF-α of clinical importance. They were measured using antibody-based method (SC: 17 patients, 30 samples; LC: 17 patients, 22 samples; Figure 5B and Figure S6C). Except for TNF-α, other molecules showed no difference between SC and LC patients, probably because most patients in this study were non-severe cases (Figure 5B). TNF-α was found to be increases in the LC group, which might be associated with the upregulated NK cells.

**Figure 5.**
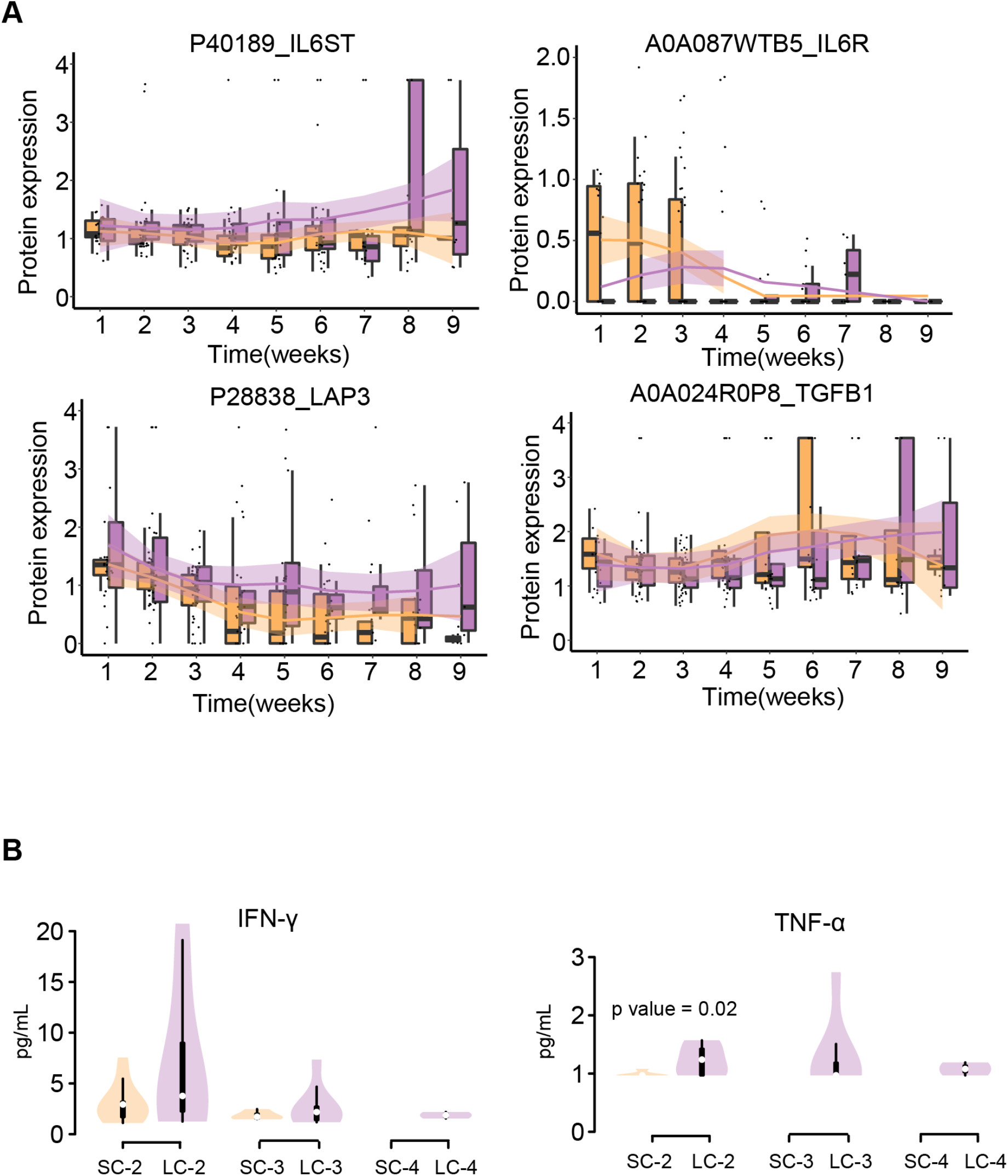
Dynamics of IL6ST, IL6R, LAP3 and TGFB1 in SC and LC patients. **(A)** Proteins detected by MS-based proteomics. **(B)** Violin plot showed that cytokines detected by antibody-based flow cytometric analysis (24 samples from 13 SC patients and 19 samples from 12 LC patients).

However, in our proteomics data, we identified two key receptors for IL-6, namely the alpha subunit (IL6R) and the beta subunit (IL6ST) of the interleukin-6 receptor. These two subunits are jointly required to mediate IL-6 signal transmission. Overall, LC patients expressed higher IL6ST than SC patients did (Figure 5). Interestingly, IL6R was upregulated in LC patients during the first four weeks followed by a decline thereafter. In SC patients, IL6R expression did not show the substantial change (Figure 5). This difference of IL6R expression trends may reflect transient activation of Th17 during the prolonged disease course. We also detected the dynamics of two proteins, transforming growth factor beta-1 protein (TGFB1) and cytosol aminopeptidase (LAP3), which are associated with Treg function. LAP3 is one of the surface markers of Treg, and TGFB1 regulated Treg and Th17 (Oida et al., 2003). Similar to IL6ST, LAP3 also kept a higher level in the LC group across the disease course than in SC patients (Figure 5). TGFB1 showed no difference between the SC and LC patients (Figure 5).

Taken together, our data showed that NK cell-mediated innate immune and Treg-mediated immunosuppression were both activated in the LC group, while CD3^-^CD56^-^ CD16^-^ cells were decreased in the LC group, suggesting suppression of B cells which may be associated with decreased antibody level.

### Predictive model for the length of disease course

The above analysis showed that SC and LC patients have different proteomic serological responses during the early phase of COVID-19 when the two groups of patients were clinically indistinguishable during the first few weeks (Figure S1). This raises the possibility of predicting disease course based on proteomic patterns during the early phase. To test this, we built a random forest model using the serum proteomic data collected during the first three weeks. We included a total of 80 serum samples as a discovery dataset (Figure 6A) which were randomly divided into two groups: a 66-sample training dataset (30 patients and 66 samples) and a validation dataset (7 patients and 14 samples). Next, a previously published cohort (39 patients and 39 samples) (Shen, 2020) was employed as an independent test dataset from a different clinical center.

**Figure 6.**
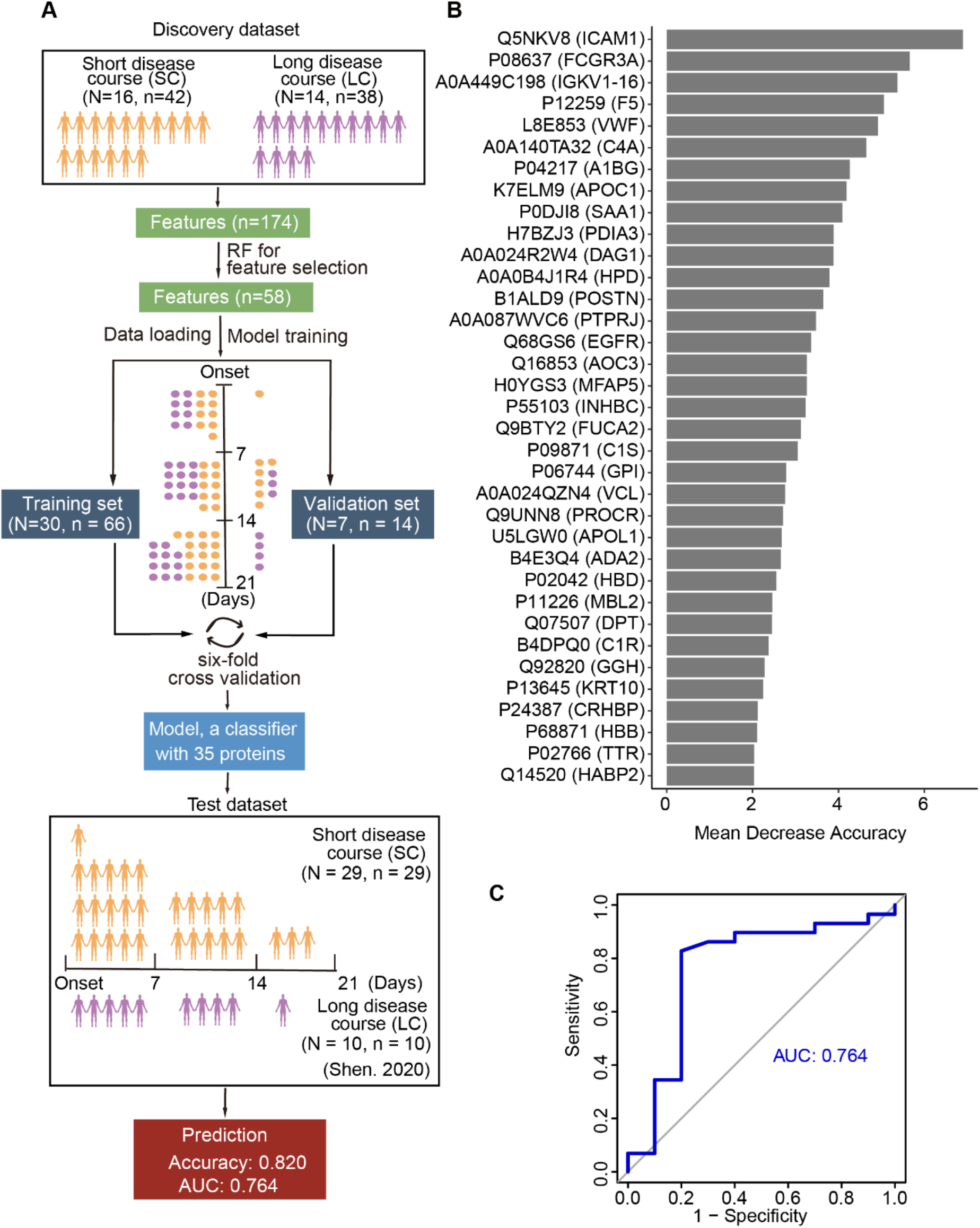
Machine learning model for predicting disease course. **(A)** Workflow for machine learning. **(B)** The top 35 key feature proteins selected by the machine learning model. **(C)** The validation of the model in an independent test dataset.

To select the most relevant proteins (referred as “feature” in machine learning), we retrieved 174 significantly differentially expressed proteins from the proteomic profiling of SC and LC patients during the first three weeks (Table S3). These proteins were also identified in the independent test dataset. Then 58 features with highest predictive accuracy were prioritized from the training set using random forest (4.6.14) (Figure 6A, Figure S7A). After a six-fold cross-validation between the training set and the validation set, a classifier with 35 feature proteins was finalized for classifying LC patients with 100% accuracy in the training set (Figure 6A). The details of the 35 key features are shown in Figure 6B. Intercellular adhesion molecule 1 (ICAM1), low affinity immunoglobulin gamma Fc region receptor 3-A (FCGR3A) and immunoglobulin kappa variable 1-16 (IGKV1-16) are the top three ranked proteins, which were significantly regulated in LC patients within the first three weeks (Figure S7B). ICAM1 mediates cell-cell contact and promotes T cell apoptosis (Starke et al., 2010). FCGR3A is involved in antibody-dependent cell-mediated cytotoxicity (ADCC). The elevation of FCGR3A is consistent with the increase of NK cells in the LC group. IGKV1-16 is a part of the variable domain of immunoglobulin light chain, and it is secreted by B cell. IGKV1-16 was down-regulated in the LC group, which might be associated with dysregulation of B cell in the LC as discussed previously.

We then applied the model to the independent test cohort from a different clinical center. The model correctly classified 32 out of 39 patients with an overall accuracy of 82% (Figure 6C). Due to biosafety issues and the emergency of this pandemic, this study is limited by the sample size. We noticed that the distribution of samples in the different disease stage is different between discovery dataset and test dataset. The numbers of serum samples of the discovery dataset are 18, 31 and 31 for the first, second and the third week, respectively. However, the numbers for the test dataset are 21, 14 and 4, showing substantially lower proportions in the second and third week. This difference was significant as measured by a two-sides Fisher’s exact test (p = 0.0004). The substantially higher number of samples collected in the first week have probably contributed to the difficulty of correct classification by this model. Indeed, the prediction accuracy was 92.9% for the samples collected in the second and third weeks, while the accuracy was only 72.2% for the samples collected in the first week. Future application and improvement of this model in larger clinical cohorts are needed.

## Conclusion

COVID-19 patients with short and long disease courses showed no significant difference in clinical symptoms, laboratory tests and response to conventional therapy. Our longitudinal sera proteomic analysis uncovered characteristic host responses in sera of patients with long disease course, including enhanced NK cell-mediated innate immune response and Treg-mediated immunosuppression at an early stage. Our data also suggested CD3^-^CD56^-^CD16^-^ cells might contribute to decreased production of S protein specific antibodies in the patients with a prolonged disease course. Our data suggest modulation of Treg cells may be potential therapeutics for these COVID-19 patients. We also showed that it is possible to predict the disease course of COVID-19 patients based on selected protein levels within the first three weeks after disease onset. The model nevertheless requires further validation in independent cohorts.

## Data Availability

All data are available in the manuscript or the supplementary materials. The proteomics data are deposited in ProteomeXchange Consortium (https://www.iprox.org/). Project ID: IPX0002170000.

## ACKNOWLEDGMENTS

This work is supported by grants from Tencent Foundation (2020), National Natural Science Foundation of China (81972492, 21904107, 81672086), Zhejiang Provincial Natural Science Foundation for Distinguished Young Scholars (LR19C050001), and Hangzhou Agriculture and Society Advancement Program (20190101A04). We thank Drs O.L. Kon, H. Qi, H. Xu, and X. Chang for helpful comments to this study, and Westlake University Supercomputer Center and biomedical research core facilities for assistance in data generation and analysis.

## AUTHOR CONTRIBUTIONS

T.G., S.T., Y.Z. and J.H. designed and supervised the project. S.T., J.H., T.M., C. H., S.L., X.X., H.L., L.W., J.D. collected the samples and clinical data. R.S., Q.X., W.G., M.L., L.Q., H.C., Q.Z., S.L., W.L., H.G., L.L., T.L., X.L., X.C., and G.R. conducted proteomic analysis. T.M., Q.X. and R.S. performed flow cytometry analysis. R.S., Q.X., W.G., T.M., C.H., Y.Z., S.T. and T.G. interpreted the data with inputs from all co-authors. R.S., Q.X., Y.Z. and T.G. wrote the manuscript with inputs from co-authors.

## METHODS

**KEY RESOURCES TABLE**

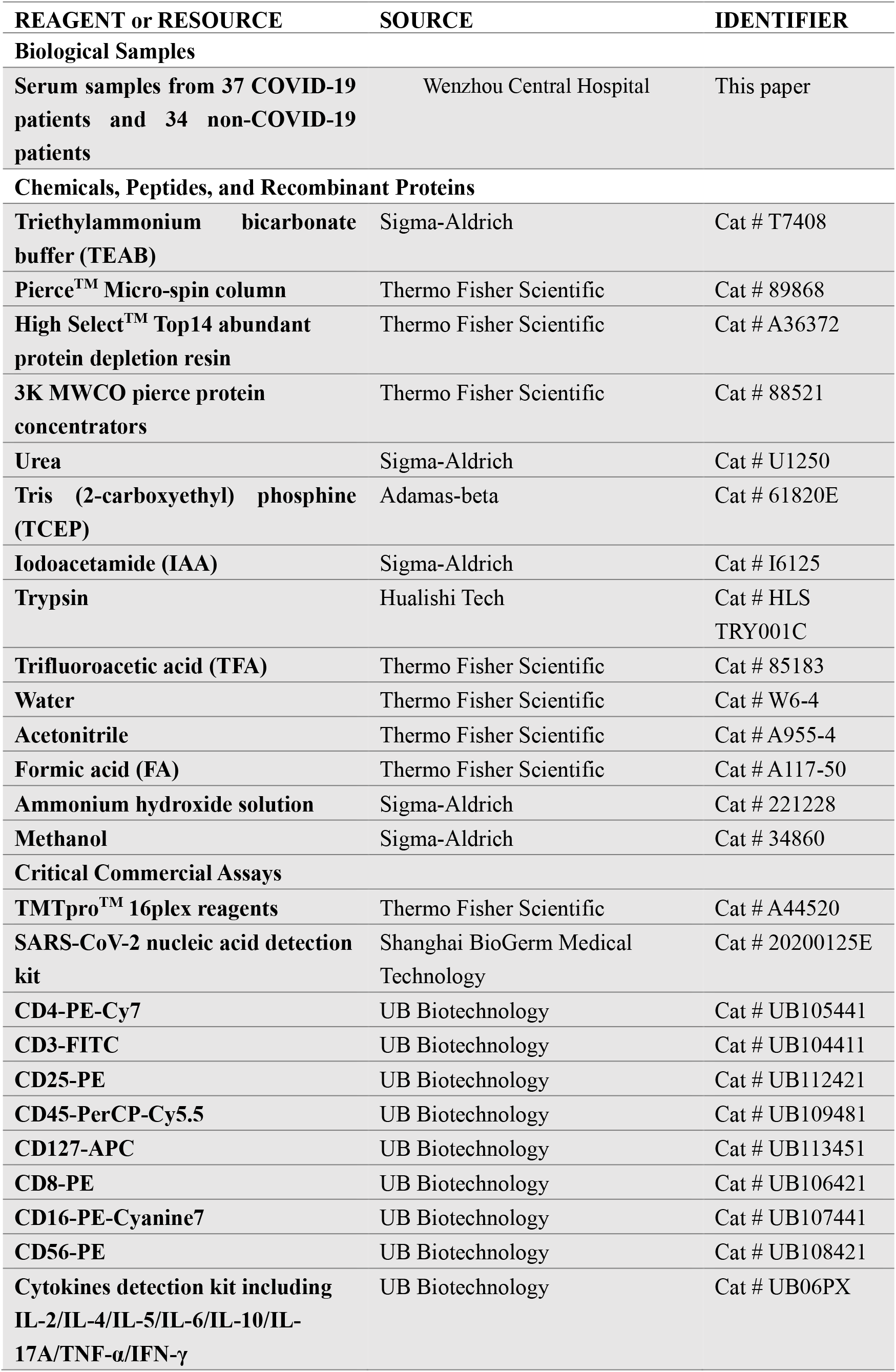
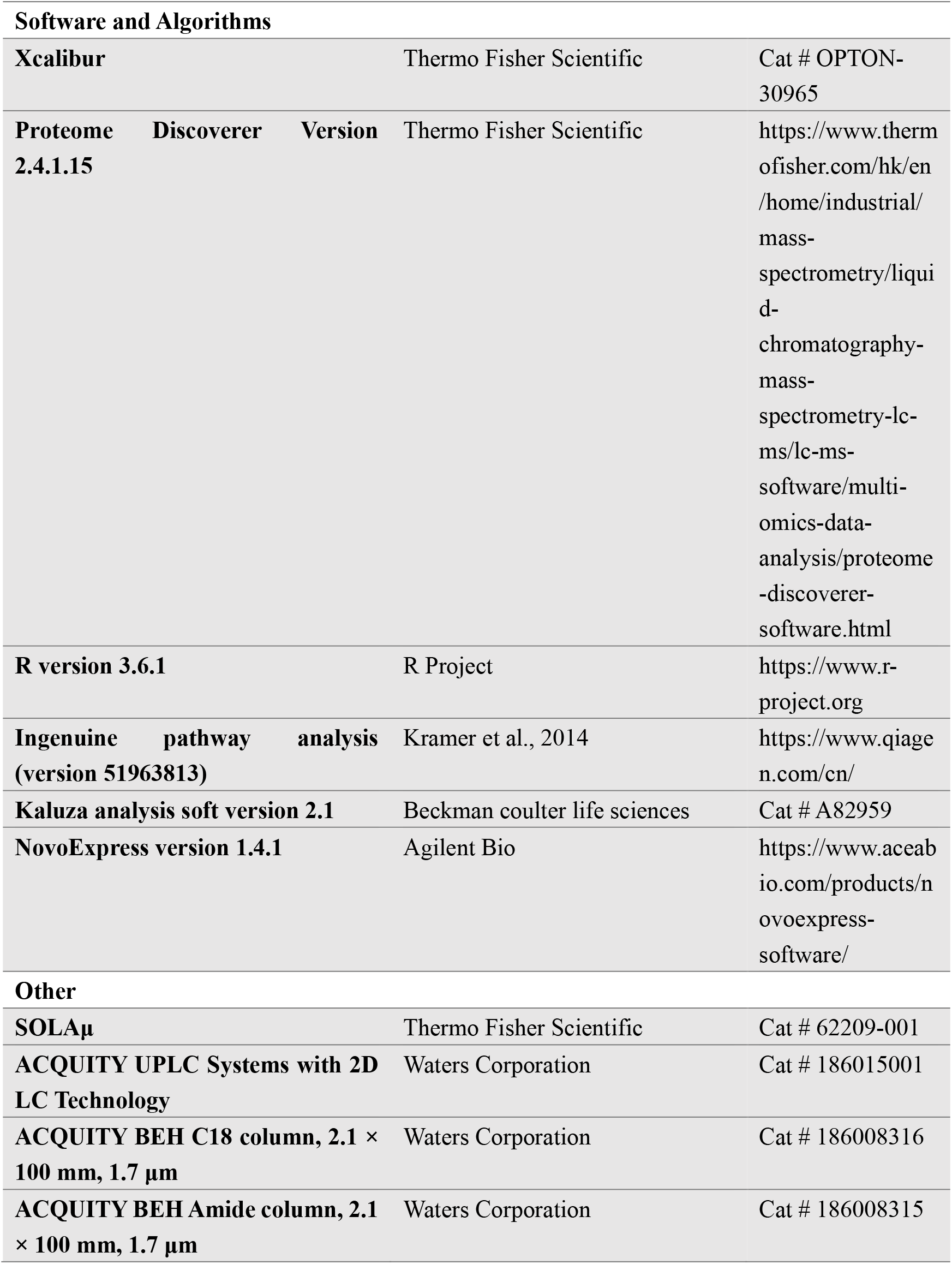

## RESOURCE AVAILABILITY

### Lead contact

Further information should be directed to and will be fulfilled by the Lead Contact Tiannan Guo.

### Materials Availability

This study did not generate new unique reagents.

### Data Availability

All data are available in the manuscript or the supplementary materials. The proteomics data are deposited in ProteomeXchange Consortium (https://www.iprox.org/). Project ID: IPX0002170000. All the data will be publicly released upon publication.

## EXPERIMENTAL MODEL AND SUBJECT DETAILS

### Patients and sera samples

We procured 72 patients in this study, including 37 COVID-19 patients whose sputa or throat swabs were tested positive for SARS-CoV-2 according to the manufacturer’s instructions (Shanghai BioGerm Medical Technology Co., LTD, Shanghai, China). According to the Chinese Government Diagnosis and Treatment Guideline (Trial 4th version) (Shen, 2020), these 37 COVID-19 patients include 35 typical cases and two severe cases. We have also procured 35 non-COVID-19 patients showing similar flu-like clinical symptoms to COVID-19 patients with negative nucleic acid testing for SARS-CoV-2. More details of these patients are provided in Figure 1A and Table S1.

We defined the disease onset as the day when the patient manifests clinical symptoms, while the disease recovery as the day when nucleic acid test of sputa or throat swab turns negative, and negative for the second test after a minimal interval of 24 hours, the same test is still negative. The disease course was thus defined as the period between disease onset and disease recovery.

Totally 224 sera samples from these patients were collected longitudinally for proteomics analysis (Figure 1B, Table S1). Sampling was performed in the early morning before diet using serum separation tubes (BD, USA). The blood was clotted for about 30 min at room temperature, and then centrifuged at 1000 g for ten min for serum sample collection. This study has been registered in the Chinese Clinical Trial Registry with an ID of ChiCTR2000031699. This study has been approved by the Ethical/Institutional Review Board of Wenzhou Central Hospital and Westlake University. Contents from patients were waived by the boards.

## METHOD DETAILS

### Proteome analysis

Serum samples were prepared as previously described (Shen, 2020). Briefly, serum samples were firstly inactivated and sterilized at 56 °C for 30 mins. For proteomic study, 4 μL of serum was used for each sample. The serum was firstly depleted of 14 high abundant serum proteins using a human affinity depletion kit (Thermo Fisher Scientific™, San Jose, USA). After depletion, the serum solution was concentrated into 50 μL through a 3k MWCO filtering unit (Thermo Fisher Scientific™, San Jose, USA) according to manufacturer. And then was mixed with 500 μL 8 M urea (Sigma) and concentrated into 50 μL. Then proteins were reduced and alkylated with 10 mM tris (2-carboxyethyl) phosphine (TCEP, Sigma) and 40 mM iodoacetamide (IAA), respectively. Proteins were submitted to two times of tryptic digestion (enzyme to protein ratio: 1:20; Hualishi Tech. Ltd, Beijing, China). The digestion was then stopped with 1% trifluoroacetic (TFA) (Thermo Fisher) to pH 2–3 to stop the reaction, and peptides were subjected to C18 (Thermo Fisher) desalting.

TMT 16-plex (Thermo Fisher) reagents were applied to label the digested peptides. The TMT labeled samples were further fractionated along a 2 hr basic pH reverse phase LC gradient using a Dinex Ultimate 3000 UHPLC (Thermo Fisher) (Li et al., 2020). LC-MS/MS analysis was performed using the Easy-nLC™ 1200 nanoLC-MS/MS system (Thermo Fisher) or a Dinex Ultimate 3000 UHPLC coupled to a Q Exactive HF or HF-X (Thermo Fisher), along a 60 min LC gradient at a flowrate of 300 nL/min as described previously (Gao et al., 2020; Shen, 2020). To reach comparable proteomic depth, the fractionated samples were combined into 30 fractions for analysis in QE-HF instruments and 26 for QE-HFX instruments.

### Database search and statistical analysis

MS data was performed using Proteome Discoverer (Version 2.4.1.15, Thermo Fisher) (Colaert et al., 2011) search engine against the human protein database downloaded from UniProt (version 02/01/2020; 164,930 sequences), with a precursor ion mass tolerance of 10 ppm and fragment ion mass tolerance of 0.02 Da. Please see previous paper (Shen, 2020) for detailed parameters of the database searching. Briefly, TMT pro-plex labels to lysine and N-terminus, and carbamidomethylation of cysteine were set as static modifications. A cut-off criterion of a q-value of 0.01, corresponding to a 1% false-discovery rate (FDR) was set for the filtered of identified peptides with highly confident peptide hits.

After filtering 80% missing rate proteins, 2700 proteins used for differential expression analysis based on |log2(FC)| and two-sided unpaired Welch’s t test. The created missing values were imputed with zero.

### Pathway analysis

For the pathway enrichment analysis, firstly, four databases including KEGG pathway, GO biological processes, Reactome gene sets and immunologic signatures were used for immune characterization analysis on the Metascape web-based platform (Zhou et al., 2019). IPA was then used to look at the pathways for differentially expressed proteins.

### Statistical analysis

Two-sided unpaired Welch’s t test was performed for each pair of comparing groups. The one-way analysis of variance (ANOVA) was used to performed among nine time points. Adjusted p values were calculated using Benjamini & Hochberg correction.

### Machine learning

The machine learning was performed using the R package randomForest (version 4.6.14) as described previously with some modifications (Shen, 2020) as described briefly in the following. The discovery dataset was divided into two parts, *i.e*. the training set and the validation set. We optimized the key random forest parameters including the cutoff values for decrease mean accuracy, cross-validation fold, and the number of trees. Input protein features were selected based on the mean decrease accuracy cutoff. For the optimized model, the minimal mean decrease accuracy of protein features was 2, the mtry was set as 13, and 800 trees were built. Six-fold cross validation was performed and this was repeated 100 times.

### Flow cytometry analysis

Peripheral blood samples from EDTA anticoagulants were incubated with mixture antibodies including CD4-PE-Cy7 (UB105441, UB Biotechnology Co., Ltd, Hangzhou, China), CD3-FITC (UB104411), CD25-PE (UB112421), CD45-PerCP-Cy5.5 (UB109481), CD127-APC (UB113451) and a kit (UB06PX) for cytokines detection including IL-2/IL-4/IL-5/IL-6/IL-10/IL-17A/TNF-α/IFN-γ for 15 min at room temperature. Double negative control and single-stain controls were prepared by normal samples, and were used to calculate a compensation matrix. Sample acquisition was performed on a Gallios (Beckman Coulter) cytometer equipped. Final analysis and graphical output were performed using NovoExpress software (Agilent Bio).

## SUPPLEMENTARY FIGURE LEGEND

**Figure S1.**
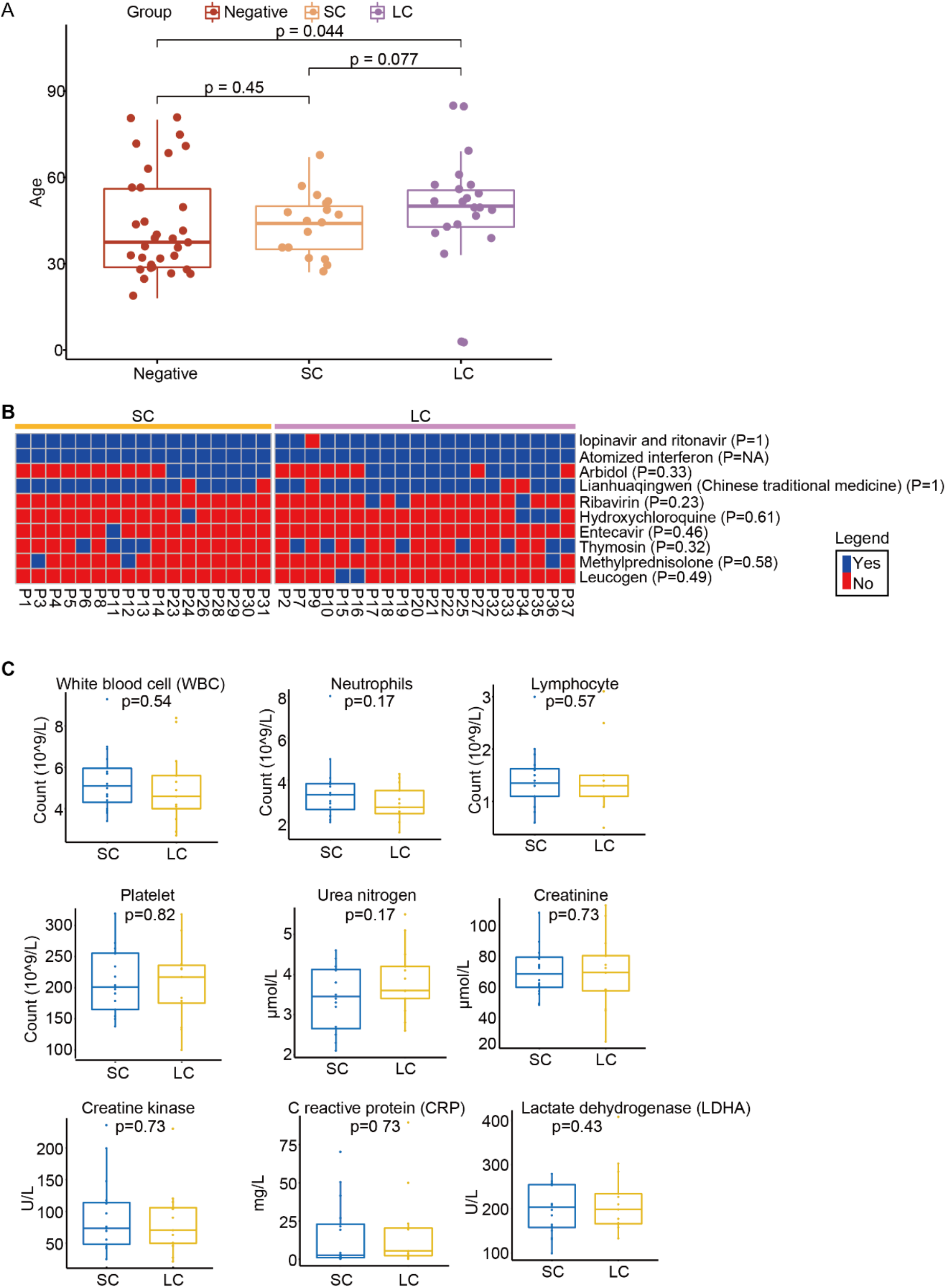
Correlation analysis between the SC and LC group based on clinical data, Related to Figure 1. **(A)** Boxplot of patient ages. Ctrl (N=35), SC (N=17) and LC(N=20). **(B)** Heatmap shows medication history in SC and LC groups. The column set on the right represents the drugs and relevant p value showing medication difference between the two groups (two-sided Fisher. test). **(C)** Comparison of blood test results between SC (N=17) and LC (N=15) patients. Boxplots display the blood biochemistry tests based on samples collected on the first day in the hospital.

**Figure S2.**
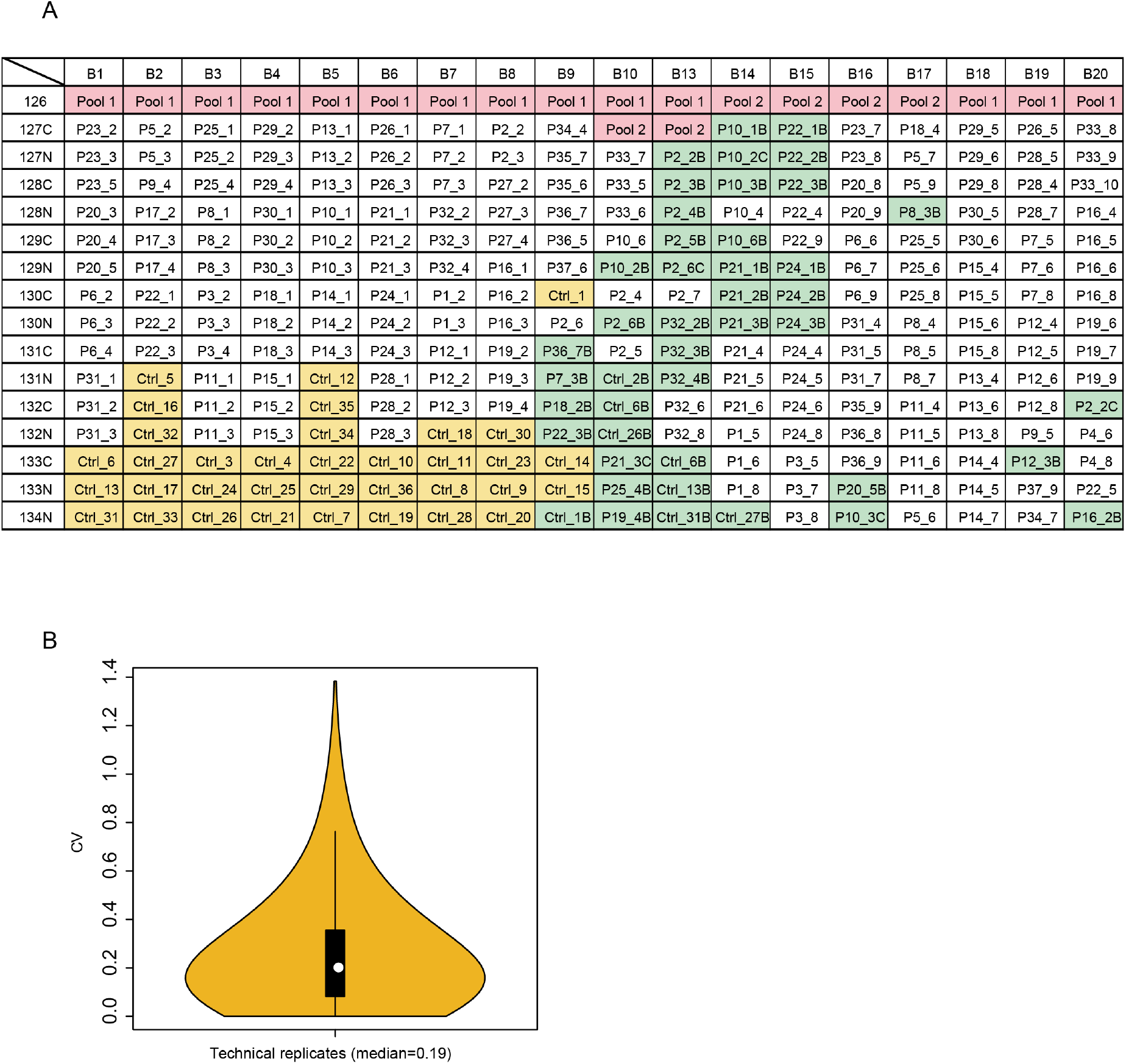
Quality assessment of proteomics data. Related to Figure 1. **(A)** Batch design of proteomic analysis. All 268 peptide samples are randomly separated into 18 batches for TMTpro labeling and MS analysis. **(B)** The median technical CV of the proteomics data is calculated by the identified proteins in 44 technical replicates.

**Figure S3.**
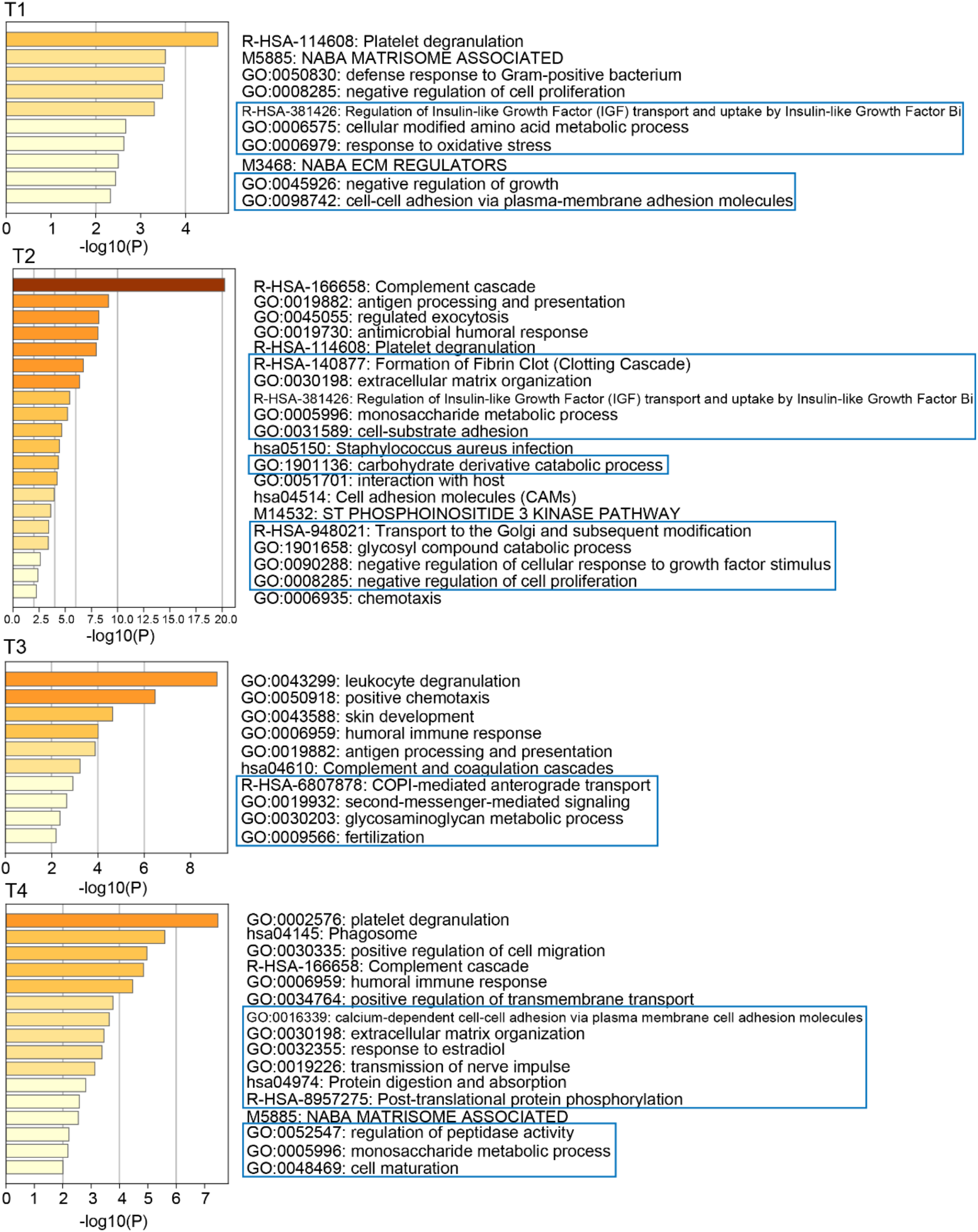
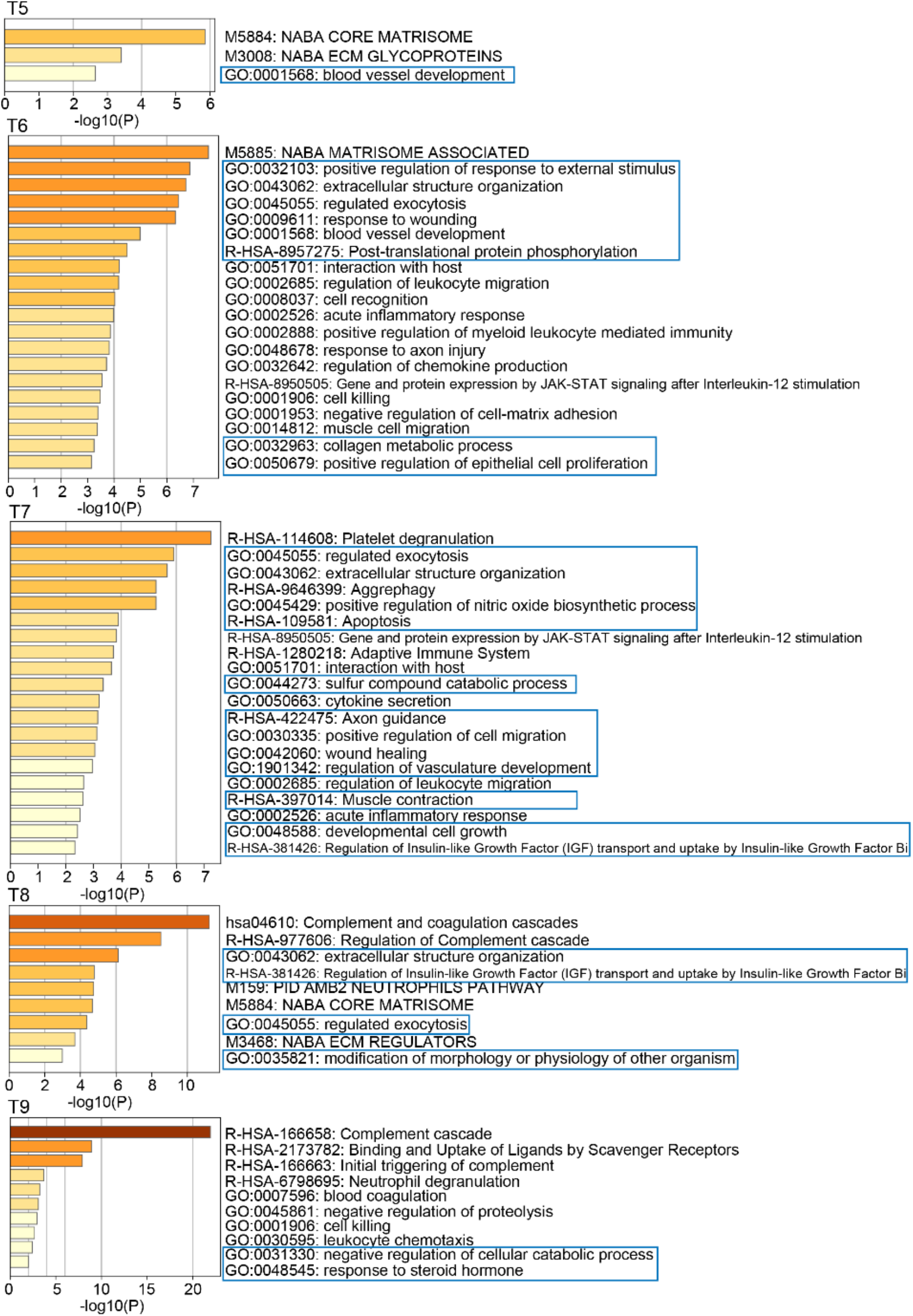
Pathway enrichment analysis at nine time points using differentially expressed proteins between the SC and LC group, Related to Figure 2. Pathways were enriched using Metascape. The pathways in the blue box are not directedly related to immunity.

**Figure S4.**
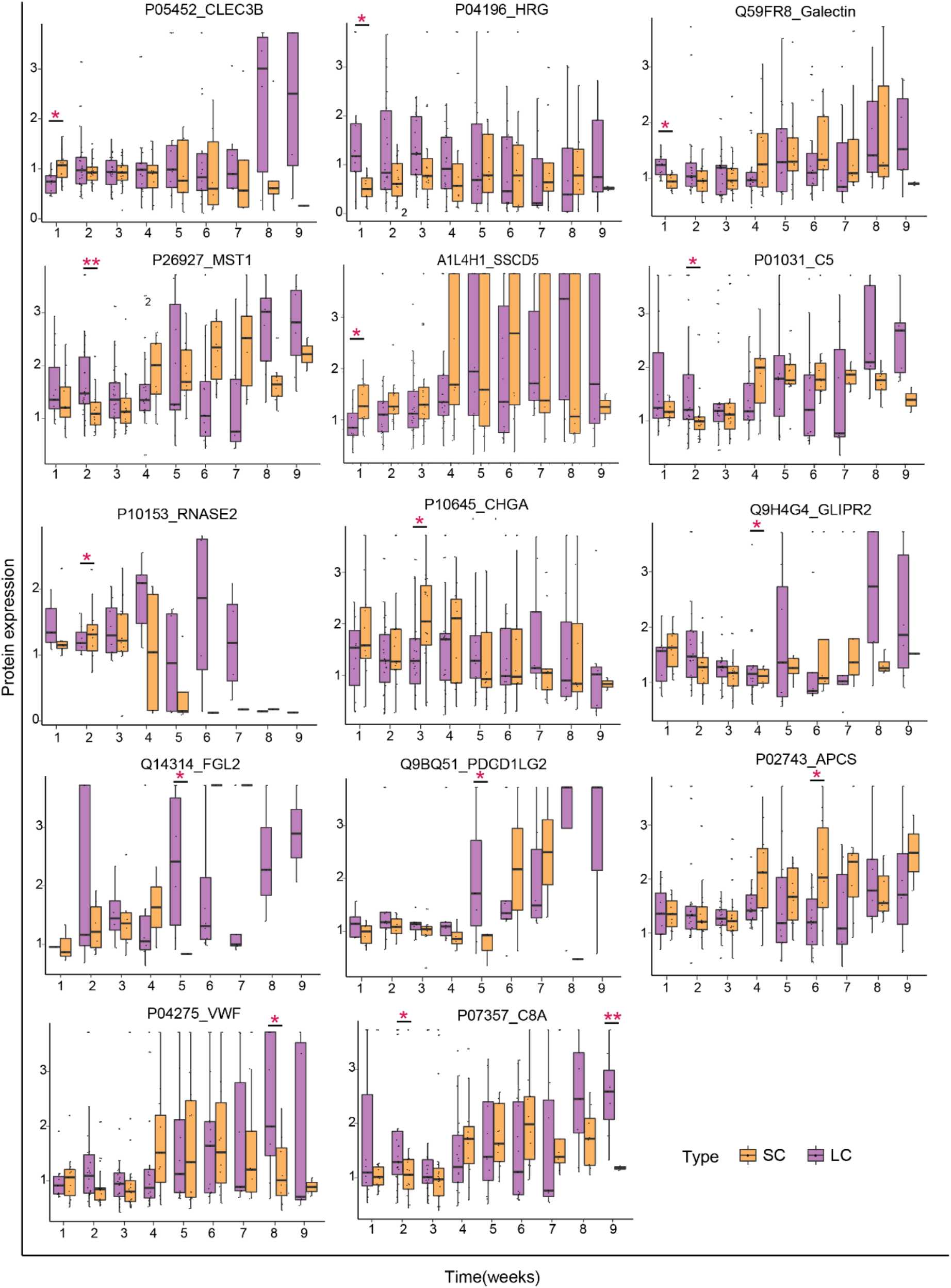
The dynamic expression of proteins across the entire disease course between SC and LC, Related to Figure 2. The x-axis shows the week index, while the y-axis denotes protein expression ratio from the TMT experiment. Pair-wise comparison of each proteins in the two patient groups was performed with student’s *t* test. *, p < 0.05; **, p < 0.01.

**Figure S5.**
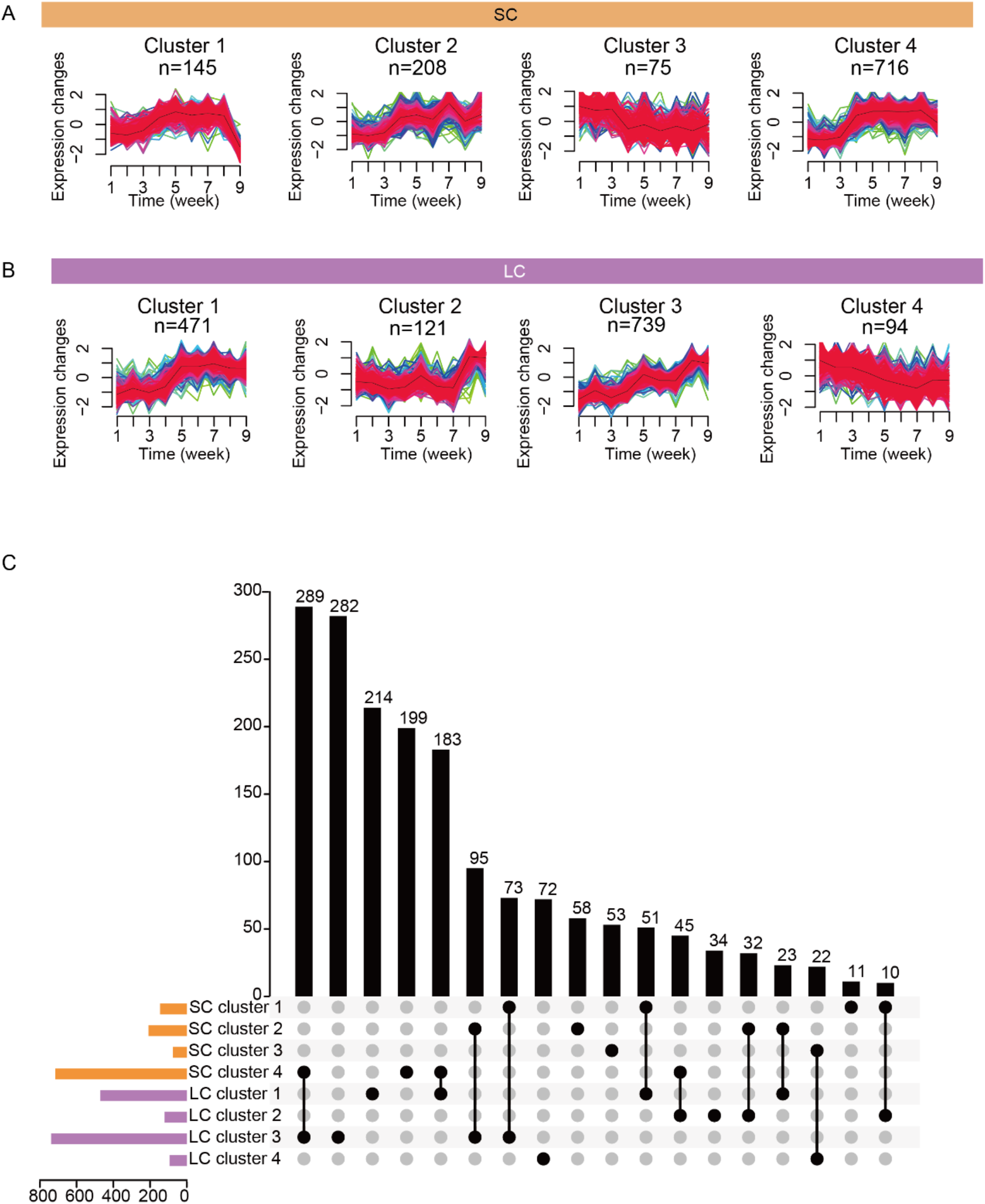
Eight clusters of proteins showing temporal dynamics by Mfuzz analysis in the SC and LC group, Related to Figure 4. Proteins detected in SC **(A)** and LC **(B)** are grouped into eight clusters with different quantitative patterns over the disease course. ANOVA analysis was performed in each cluster (the cutoff of the adjusted p value (BH adjust) was 0.05). **(C)** Upset plots shows the overlapping protein clusters selected by Mfuzz.

**Figure S6.**
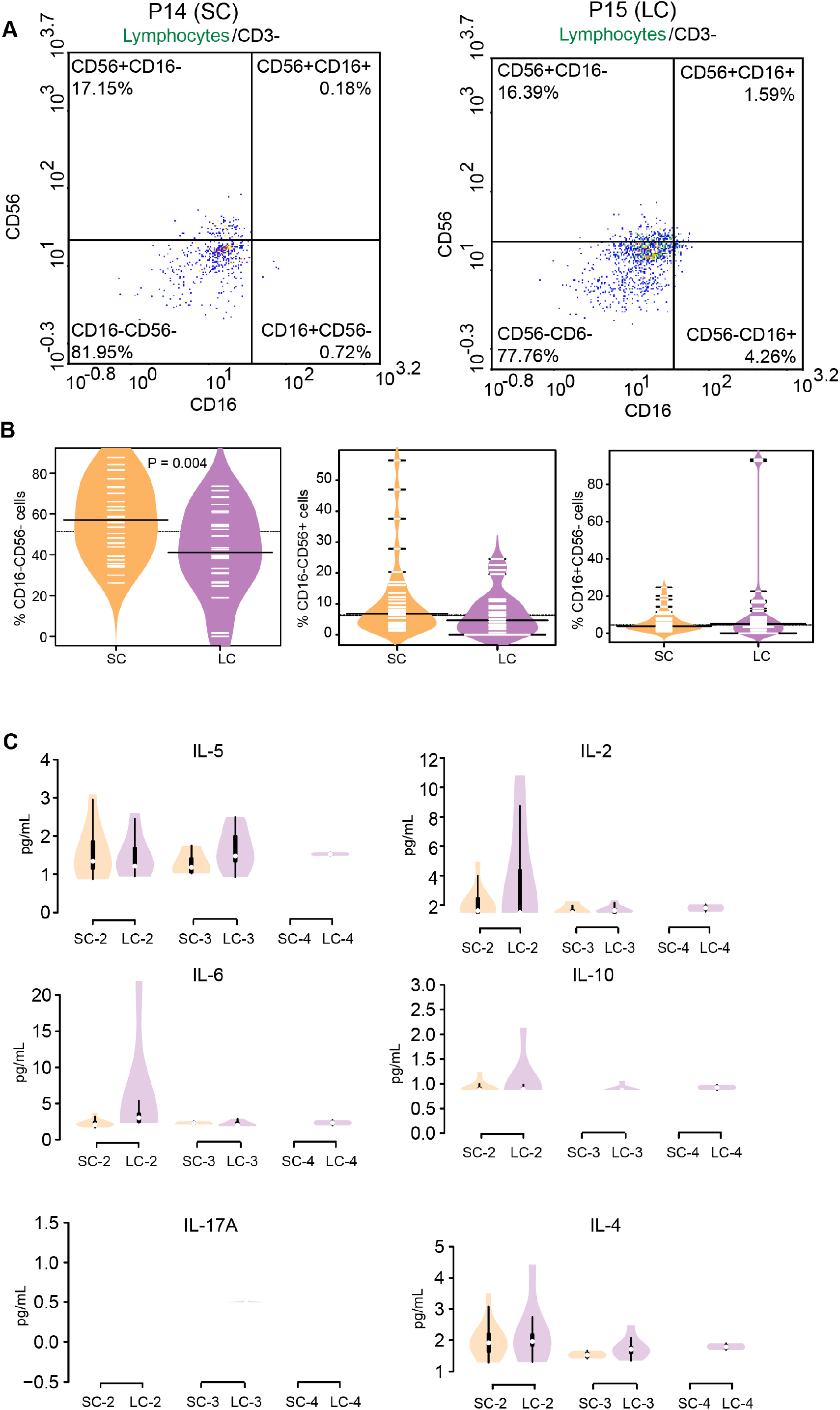
Flow cytometric analysis of lymphocytes, CD3-cells, Related to Figure 4. 22 samples from 17 LC patients and 30 samples from 17 SC patients were measured and analyzed. **(A)** Results of lymphocytes, CD3-cells, i.e. P14 (SC) and P15 (LC). **(B)** Bean plots show the comparison of cell populations between patient groups. **(C)** Violin plots show cytokines detected by antibody-based flow cytometric analysis (24 samples from 13 SC patients and 19 samples from 12 LC patients). The y-axis shows the intensity of each cytokine.

**Figure S7.**
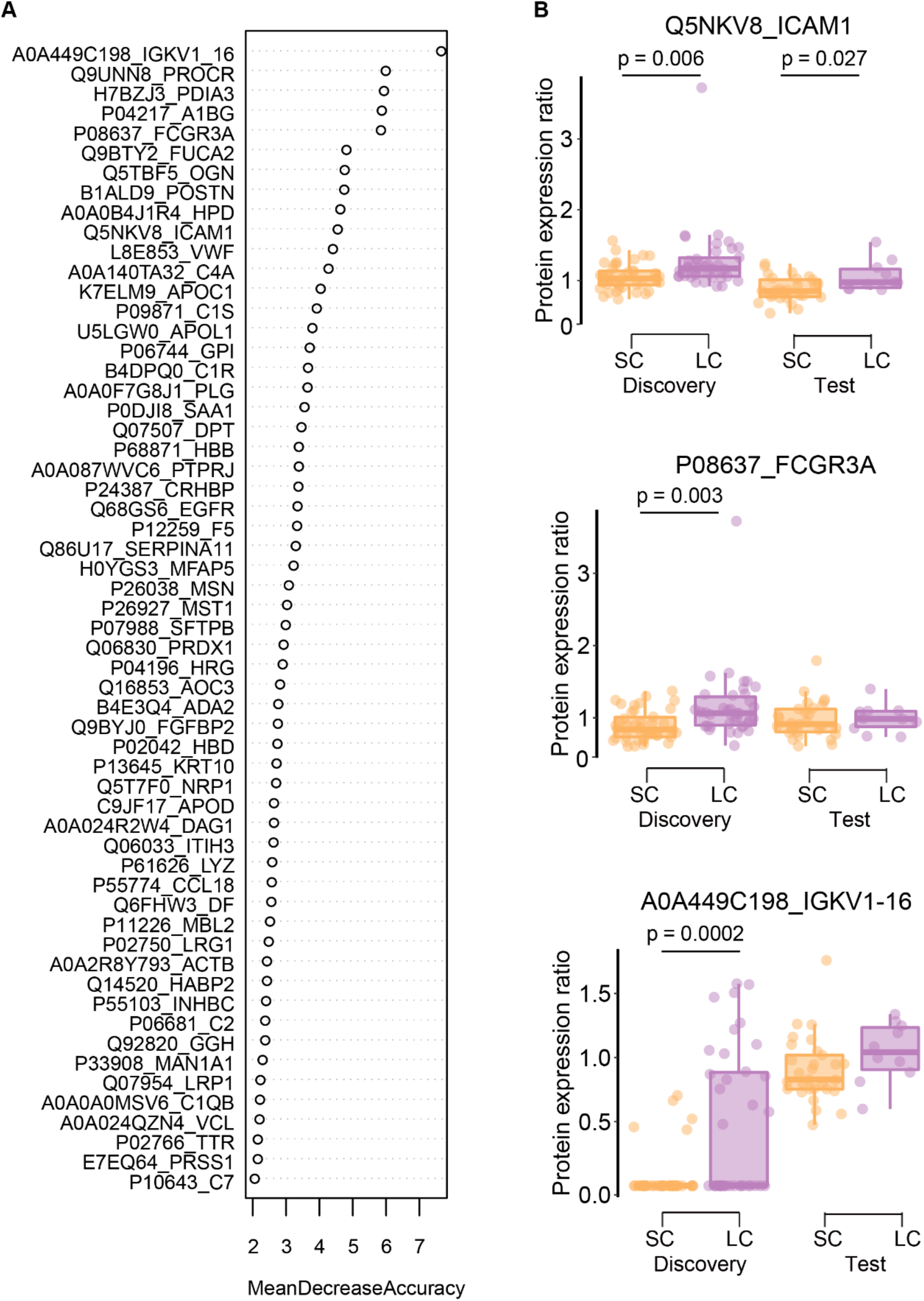
58 key feature proteins and the expression of top three proteins, ICAM1, FCGR3A and IGKV1-16. Related to Figure 6. **(A)** 58 features ranking according to mean decrease accuracy. **(B)** Protein expression of ICAM1, FCGR3A and IGKV1-16 between SC and LC groups in discovery dataset test dataset, respectively.

